# NGFR as a biomarker and actionable target in cisplatin-based chemoradiotherapy–resistant HNSCC

**DOI:** 10.64898/2025.12.25.25342978

**Authors:** Juan García-Agulló, Vanesa Santos, Yoelsis Garcia-Mayea, Beatriz de Luxan-Delgado, Marina Bataller, Matilde Esther Lleonart, Juan P. Rodrigo, Juana M. García-Pedrero, Mónica Álvarez-Fernández, Héctor Peinado

**Author notes:** Corresponding author: Héctor Peinado, PhD, Department of Immunology and Oncology, Centro Nacional de Biotecnología (CNB-CSIC), C. Darwin, 3, 28049 Madrid, Spain. **Novelty and Impact:** We found that the nerve growth factor receptor (NGFR) mediates resistance to cisplatin-based therapy in head and neck squamous cell carcinoma (HNSCC). In patient treated with radiotherapy and/or chemoradiotherapy, NGFR expression was associated with tumor recurrence. Moreover, NGFR targeting in the cisplatin-resistant human HNSCC cell line Detroit 562 restored sensitivity to cisplatin. Together, these findings support NGFR as a candidate biomarker for stratifying patients at risk of cisplatin resistance and as a promising therapeutic target in cisplatin-resistant disease models.

## Abstract

Head and neck squamous cell carcinoma (HNSCC) is an aggressive malignancy with high mortality rates, often exhibiting resistance to conventional treatments such as radiotherapy (RT) or a combination of chemotherapy and radiotherapy (CRT). The nerve growth factor receptor (NGFR, also known as p75NTR or CD271) is a well-established cancer stem cell marker in melanoma, where it has been linked to resistance to multiple therapies. In HNSCC, NGFR has been reported as a poor prognostic marker, with its overexpression associated with disease progression. However, its contribution to therapy resistance in HNSCC remains unknown. Here, we show in a cohort of RT/CRT-treated patients that NGFR expression identifies individuals with poor prognosis and increased risk of recurrence following standard RT/CRT. Moreover, we found that NGFR is upregulated in the human Detroit 562 cisplatin (CDDP)-resistant HNSCC cell line *in vitro* and *in vivo*. Functional studies demonstrated that genetic knock out of NGFR in these cisplatin-resistant cells restored sensitivity to CDDP *in vivo*. These results indicate that NGFR contributes to cisplatin resistance in HNSCC. NGFR is upregulated in tumors from patients with poorer prognosis and an increased risk of recurrence after standard radiotherapy and/or RT/CRT, as well as in cisplatin-resistant models. Altogether, our findings open the way to consider NGFR as a new potential therapeutic target to overcome or mitigate cisplatin resistance in HNSCC.

## Introduction

Head and neck squamous cell carcinoma (HNSCC) ranks as the sixth most frequently diagnosed cancer worldwide and is associated with high mortality [1]. These tumors are highly heterogeneous in anatomical site, etiology, and clinical presentation, arising from the mucosal lining of four anatomical regions: the sinonasal cavity, oral cavity, pharynx, and larynx [1]. According to GLOBOCAN 2022, HNSCC accounted for ∼950,000 new cases worldwide, representing ∼4.74% of all cancer diagnoses, and ∼480,000 deaths (4.95% of the total) [2]. Moreover, HNSCC incidence is expected to rise, particularly in Europe and the United States, largely due to the growing prevalence of human papillomavirus (HPV) infections [3].

HPV-negative HNSCC accounts for the majority of cases and are generally associated with a poor prognosis, with limited improvement in survival in recent decades [4]. While early-stage tumors have 5-year survival rates of ∼70–90%, most patients are diagnosed with locally advanced or metastatic disease, in which survival is markedly reduced and current therapies frequently fail [5]. For locally advanced disease, the standard of care is concurrent chemoradiotherapy (CRT), typically with high-dose cisplatin in combination with intensity-modulated radiation therapy (IMRT) [5–7]. Cisplatin-based CRT offers the greatest survival benefit when tolerated; however, its significant toxicity limits its use in many patients, especially in older individuals or those with comorbidities [5, 8, 9]. Ultimately, resistance to cisplatin-based CRT is one of the major therapeutic challenges in advanced HPV-negative HNSCC. Identifying the underlying mechanisms of resistance and developing strategies to target them is essential for improving survival and represents an unmet clinical need.

The nerve growth factor receptor (NGFR), also known as p75^NTR^ or CD271, is a low-affinity transmembrane receptor for all neurotrophins and proneurotrophins [10, 11]. NGFR has emerged as a critical player in cancer. In melanoma, NGFR is a well-established cancer stem cell (CSC) marker [12], where its expression identifies tumor cells with high tumor-initiating potential, stem-like features, and pro-metastatic ability [13–15]. Importantly, NGFR expression in melanoma is also associated with resistance to BRAF inhibitors (BRAFi) and MEK inhibitors (MEKi) [16–19]. Additionally, in melanoma NGFR has also been implicated in chemotherapy resistance, through suppression of the p53 axis, which confers resistance to cisplatin and doxorubicin [20]. These findings reinforce the concept of NGFR as a central regulator in cancer cell plasticity and therapeutic escape, traits commonly associated with cancer stem cell behaviour (CSCs) [21].

In HNSCC, NGFR expression in tumors has been correlated with poor prognosis and reduced survival [22]. Initial functional analyses using hypopharyngeal models demonstrated that NGFR^+^ cancer cells possess high tumor-initiating capacity, and co-expression analyses linked NGFR to markers of stemness (e.g., Nanog), regulators of invasion and extracellular matrix (ECM) remodeling (e.g., MMPs), and chemotherapy resistance (e.g., ABC transporters) [23]. Moreover, NGFR was shown to promote invasion via regulation of ESM1, a key ECM component [24], and to directly regulate Snai2/Slug, a master regulator of epithelial–mesenchymal transition (EMT), thereby enhancing invasiveness and metastatic potential [25]. However, despite these associations, the specific contribution of NGFR to therapy resistance in HNSCC and particularly to cisplatin-based CRT remains insufficiently defined.

In this study, we studied the relevance of NGFR in radiotherapy/chemoradiotherapy (RT/CRT) resistance in a cohort of HPV-negative HNSCC-treated patients. We found that NGFR expression in tumor cells is a marker of post-treatment recurrence and predicts worse survival independently of clinical variables such as stage. Furthermore, we show that NGFR is upregulated in the human Detroit 562 cisplatin (CDDP)-resistant model and that NGFR knockout restores treatment sensitivity *in vivo*, reducing primary tumor growth and improving survival. These findings highlight NGFR as both a prognostic biomarker and a potential therapeutic target in cisplatin-resistant HNSCC.

## Results

### NGFR is expressed heterogeneously in a cohort of HNSCC patients

To investigate the relevance of NGFR in RT/CRT resistance in HNSCC, we analyzed NGFR expression in a cohort of 52 HPV-negative patients since these tumors are associated with poor prognosis and therapeutic resistance in HNSCC (**Supplementary Table 1**) [4, 5]. We found that NGFR expression was heterogeneous across the cohort: a few cases showed no detectable expression (NGFR-neg: n = 6) (**Fig. 1A**), the majority displayed low expression (NGFR-low: n = 27) (**Fig. 1B**), and a subset exhibited high expression levels (NGFR-high: n = 19) (**Fig. 1C**).

**Figure 1.**
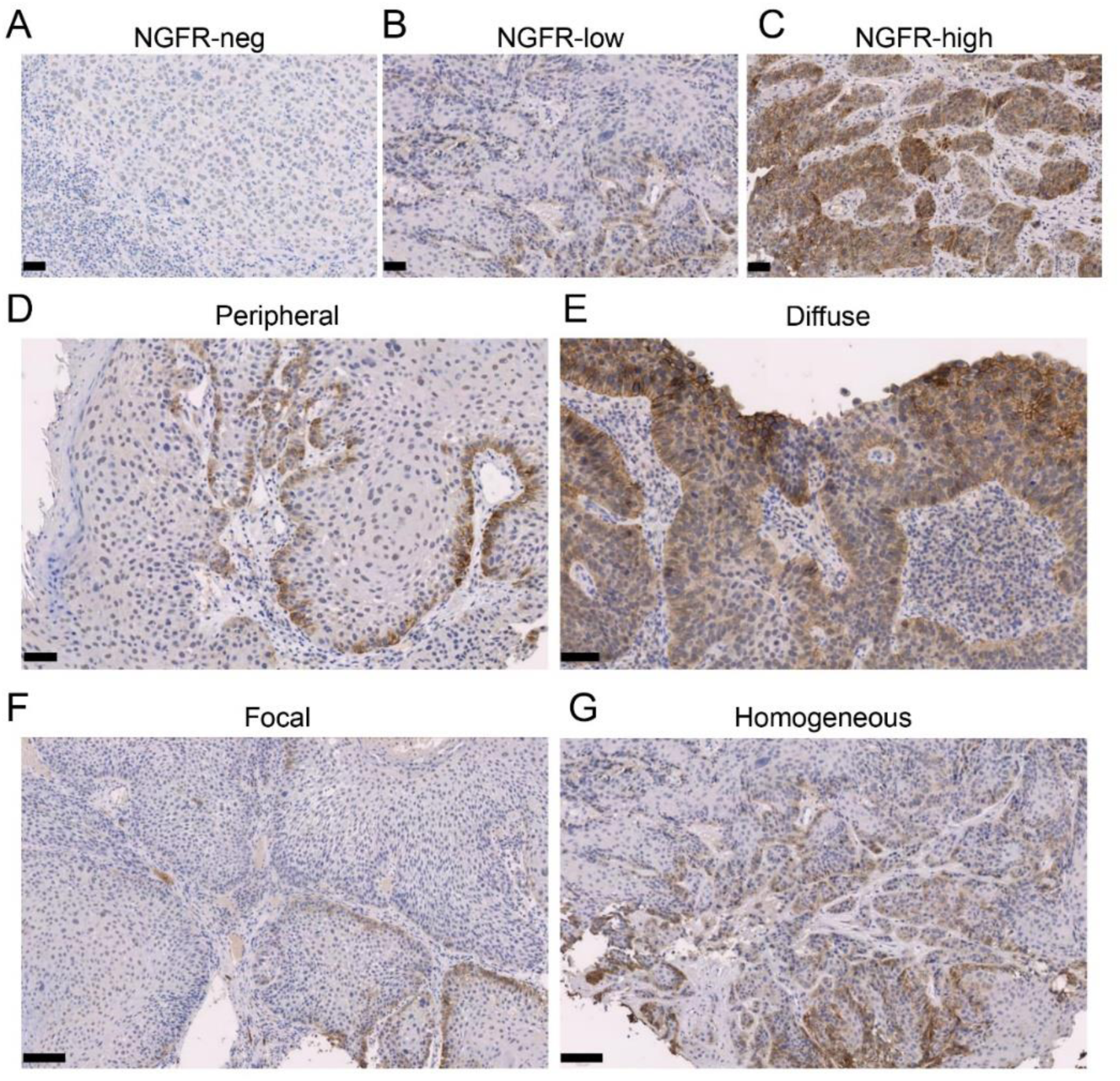
NGFR is expressed heterogeneously in HNSCC human tumors. (**A–C**) Representative images of NGFR staining intensity: negative (**A**), low (**B**), and high (**C**). (**D, E**) Distinct spatial staining patterns of NGFR expression: peripheral (**D**) and diffuse (**E**). (**F, G**) Tissue distribution patterns of NGFR expression: focal (**F**) and homogeneous (**G**). Scale bars: 50 μm (**A-E**) and 100 μm (**F, G**).

Among NGFR^+^ cases, we observed heterogeneity in expression patterns. Therefore, NGFR immunostaining was classified into two staining patterns. Some tumors displayed a peripheral pattern, with NGFR expression restricted to the tumor-stroma interface, as previously described [22] (n = 11) (**Fig. 1D**). However, most tumors showed a diffuse staining pattern, with widespread NGFR expression throughout the tumor without clear localization (n = 34) (**Fig. 1E**). In some cases, NGFR expression appeared focal, restricted to specific tumor regions (n = 13) (**Fig. 1F**), while in most cases, expression was homogeneous across the tumor biopsy (n = 33) (**Fig. 1G**).

### NGFR expression is associated with poor prognosis in HNSCC patients

We next assessed the association between NGFR expression and survival outcomes. High NGFR expression was associated with poorer recurrence-free survival (RFS) in the HPV-negative oropharyngeal subgroup (N = 46; log-rank, p = 0.0363) (**Fig. 2A**). Incorporating the staining pattern into a combined NGFR score further improved risk stratification in this subgroup, with patients whose tumors showed both high NGFR intensity and a diffuse staining pattern showing the worst RFS compared with NGFR-negative/low cases (N = 45; log-rank, p = 0.0031) (**Fig. 2B**). Importantly, these associations were maintained when the analysis was restricted to HPV-negative oropharyngeal tumors treated with cisplatin (CDDP), where high NGFR intensity (N = 30; log-rank, p = 0.0194) and the high/diffuse combined NGFR score (N = 30; log-rank, p = 0.0107) were both associated with reduced RFS (**Fig. 2C, D**). Similar trends were observed when all HPV-negative cases were considered. For RFS, high NGFR intensity was associated with poorer outcome, although this did not reach statistical significance (**Supplementary Fig. 1A**; N = 51; log-rank, p = 0.1070), whereas the combined NGFR score provided a clearer separation between groups (**Supplementary Fig. 1B**; N = 50; log-rank, p = 0.0272). For DSS, analogous patterns were seen for both NGFR intensity (**Supplementary Fig. 1C**; N = 52; log-rank, p = 0.2167) and the combined score (**Supplementary Fig. 1D**; N = 51; log-rank, p = 0.1089), although these comparisons did not reach statistical significance.

**Figure 2.**
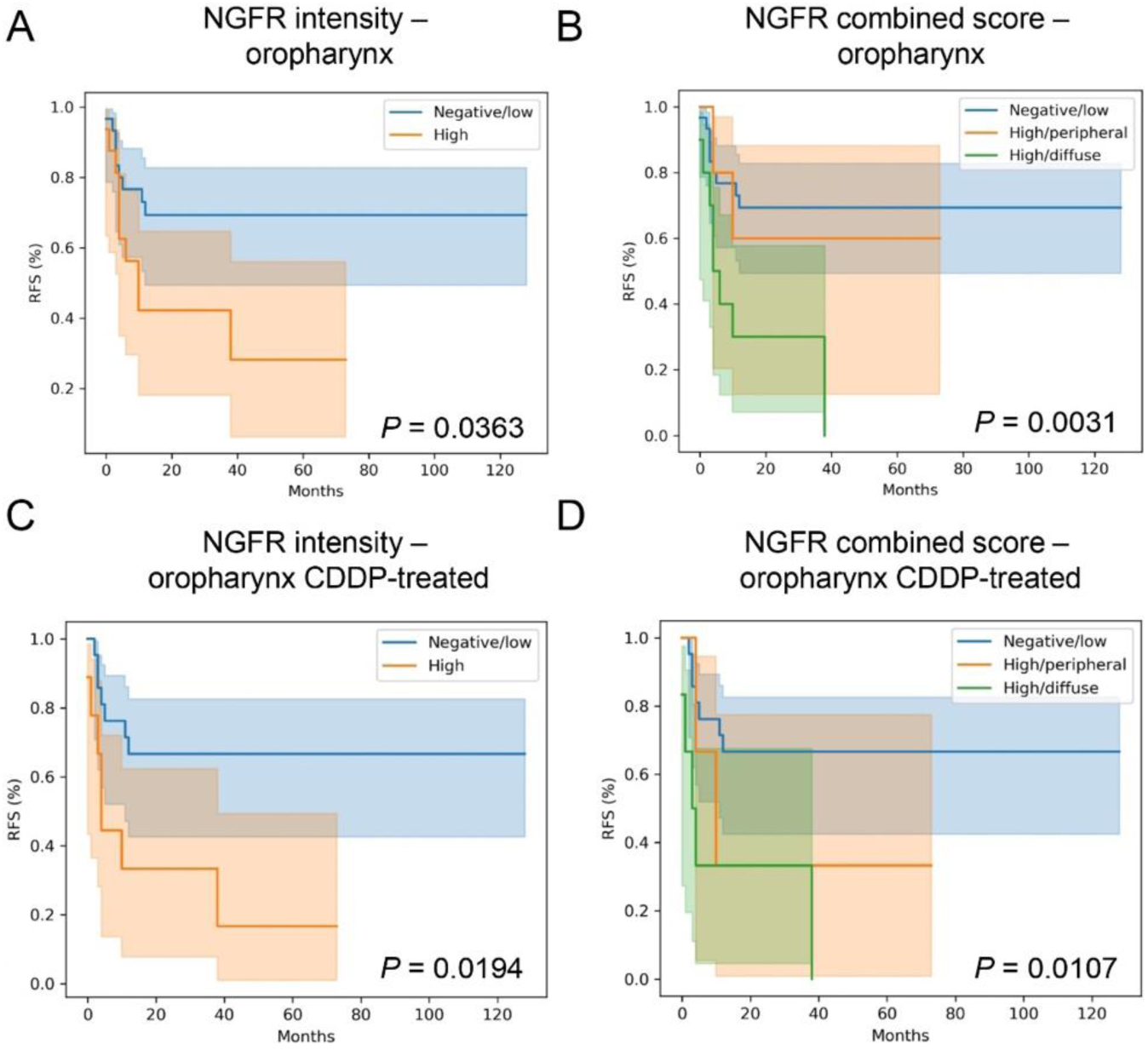
High NGFR expression is associated with poor recurrence-free survival in HPV-negative oropharyngeal HNSCC. (**A, B**) Kaplan–Meier curves for recurrence-free survival (RFS) in HPV-negative oropharyngeal tumors, stratified according to NGFR intensity (**A**) or the combined NGFR score (**B**). NGFR intensity was stratified as high versus negative/low expression. The combined NGFR score was calculated as the product of intensity (0 = negative/low, 1 = high) and staining pattern (1 = peripheral, 2 = diffuse). (**C, D**) Kaplan–Meier curves for RFS in HPV-negative oropharyngeal tumors treated with cisplatin (CDDP), stratified by NGFR intensity (**C**) or the combined NGFR score (**D**). P-values were calculated using the log-rank test. In panels **B** and **D**, comparisons are shown between negative/low and high/diffuse groups.

Additionally, we evaluated whether the prognostic value of NGFR in this cohort. To address this, we performed multivariable Cox proportional hazards analyses for both RFS and DSS. In HPV-negative oropharyngeal tumors, high NGFR intensity remained an independent predictor of shorter RFS after adjustment for these covariates (**Fig. 3A**), and this effect was even more pronounced for tumors with a high/diffuse NGFR pattern in the combined score (**Fig. 3B**). Importantly, the association between NGFR and poor RFS was maintained when restricting the analysis to HPV-negative oropharyngeal tumors treated with CDDP, both when NGFR was modeled as intensity alone (**Fig. 3C**) and as a combined high/diffuse score (**Fig. 3D**). Similar trends were observed for DSS in these subgroups and, more broadly, for both RFS and DSS in the entire HPV-negative cohort (**Supplementary Table 2**). Taken together, these multivariable analyses support NGFR, particularly high/diffuse expression, as an independent adverse prognostic marker in HPV-negative HNSCC. Importantly, we observed a stronger effect when the population was enriched for oropharyngeal tumors and for patients treated with CDDP.

**Figure 3.**
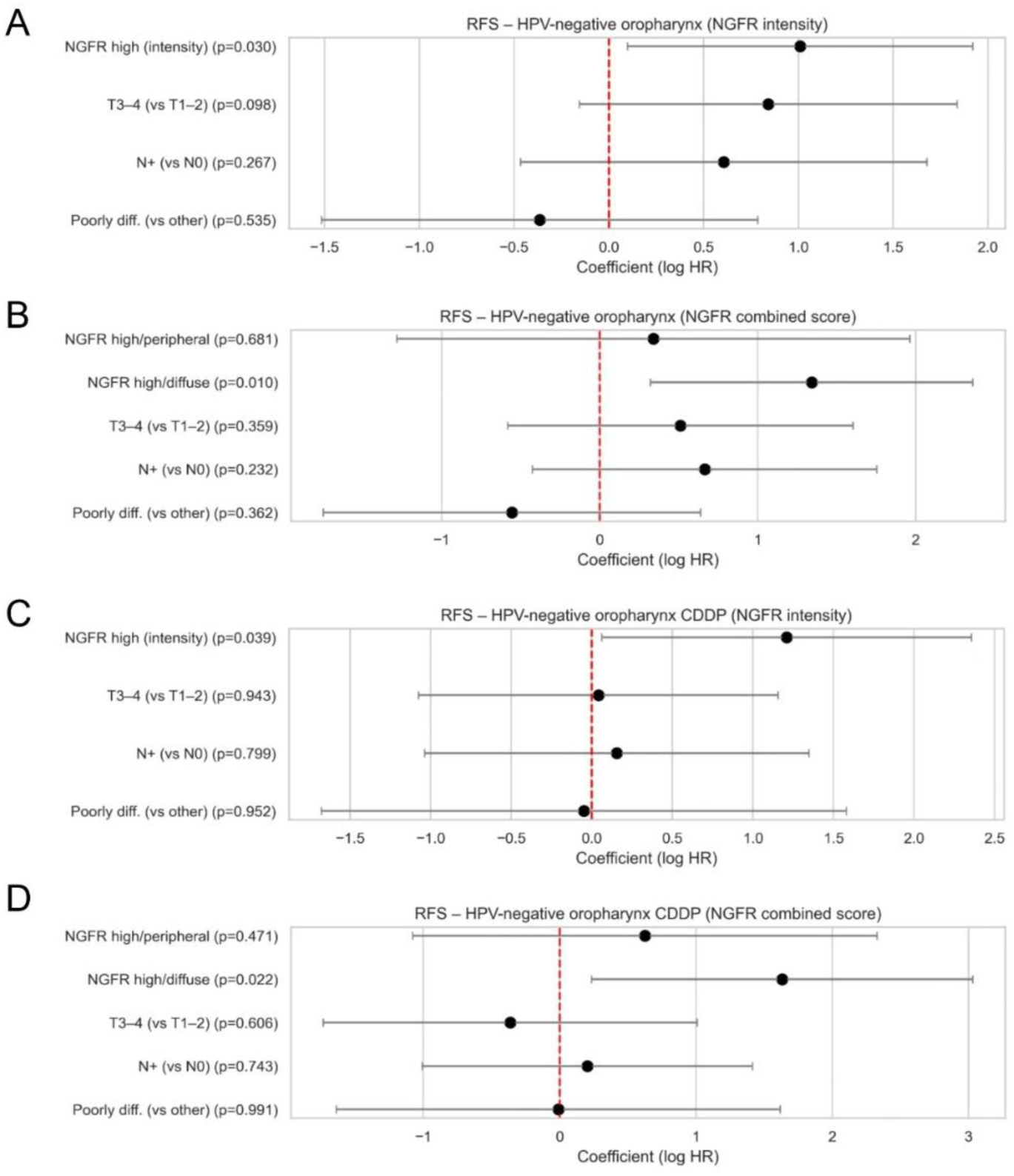
NGFR is an independent prognostic marker in an HPV-negative oropharyngeal HNSCC cohort. (**A, B**) Forest plots showing hazard ratios for recurrence-free survival (RFS) in HPV-negative oropharyngeal tumors, modeled according to NGFR intensity (**A**) or the NGFR combined score (**B**), together with T stage (T3–4 vs T1–2), nodal status (N+ vs N0) and histological grade (poorly differentiated vs other). NGFR intensity was stratified as high versus negative/low expression. The combined NGFR score was calculated as the product of intensity (0 = negative/low, 1 = high) and staining pattern (1 = peripheral, 2 = diffuse), with negative/low cases used as the reference and high/peripheral and high/diffuse categories entered as separate covariates. (**C, D**) Forest plots showing multivariable Cox models for RFS restricted to HPV-negative oropharyngeal tumors treated with CDDP, using NGFR intensity (**C**) or the combined NGFR score (**D**) together with the same clinical covariates. In all panels, points represent regression coefficients (log hazard ratios) and horizontal bars indicate 95% confidence intervals. P-values were calculated using Wald tests from Cox proportional hazards regression models.

### NGFR expression is associated with tumor recurrence in HNSCC patients

We next evaluated the association between NGFR expression levels and post-treatment recurrence in HNSCC patients. Notably, NGFR intensity was significantly associated with recurrence in HPV-negative cases (**Table 1** and **Fig. 4A, B**). Local recurrence was not observed in NGFR-negative patients, but occurred in 2 of 27 NGFR-low patients (7.4%) and 10 of 19 NGFR-high patients (52.6%). A similar pattern was observed for overall tumor recurrence, which included local, regional, and metastatic events but excluded second primary tumors, with recurrence observed in 1 of 6 NGFR-negative patients (16.7%), 10 of 27 NGFR-low patients (37.0%), and 12 of 19 NGFR-high patients (63.2%). Representative images of RT/CRT-sensitive and recurrent patients are shown in **Fig. 4C**.

**Figure 4.**
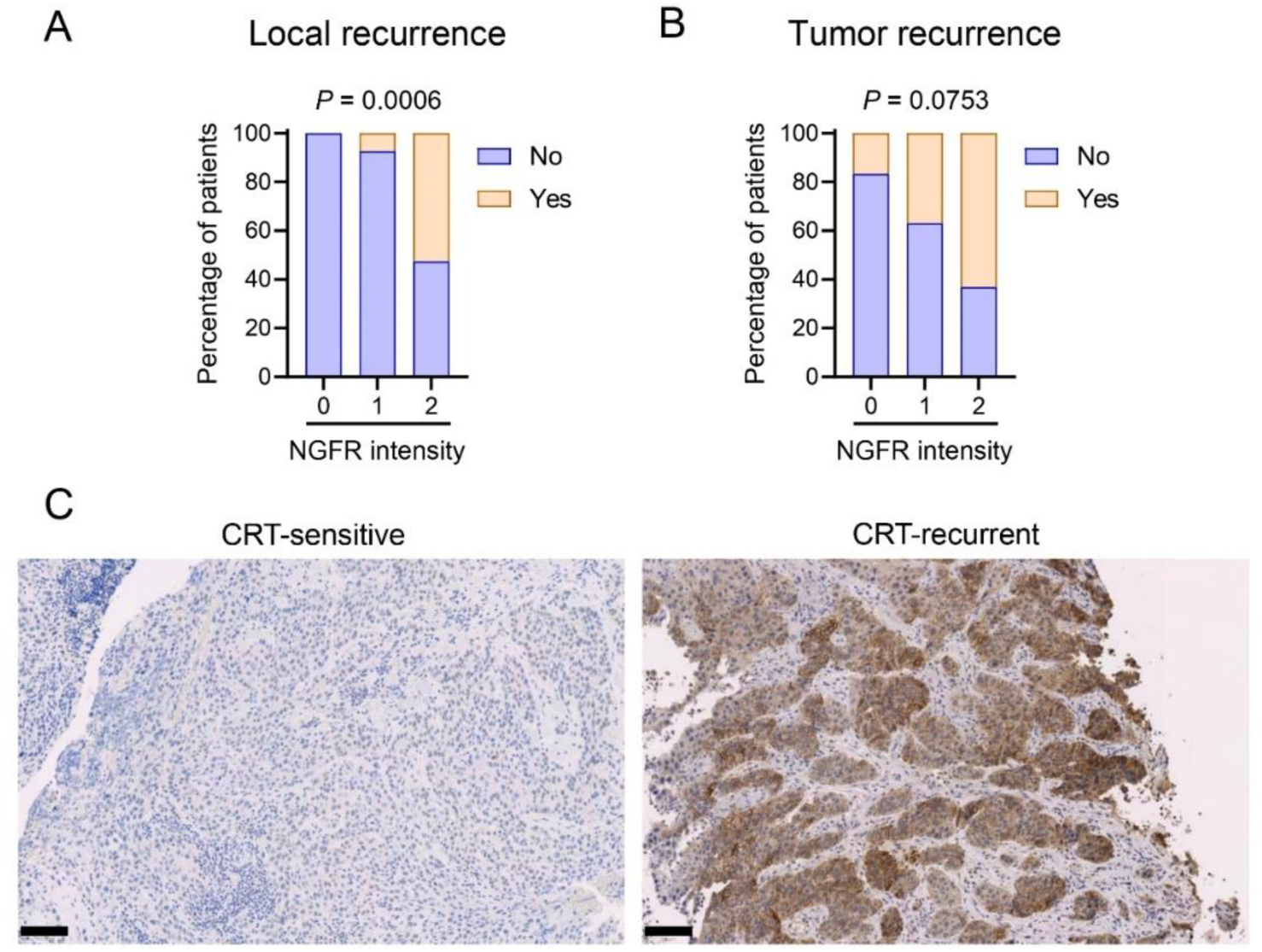
High NGFR expression is strongly associated with recurrence in HPV-negative HNSCC. (**A, B**) Bar plots of recurrence events in HPV-negative cases classified by NGFR expression intensity, categorized as negative (0), low (1), and high (2). Local recurrence is shown in panel **A** and tumor recurrence in panel **B**. Tumor recurrence includes local, regional, and metastatic events, but excludes second primary tumors. Percentages are shown in parentheses and refer to the proportion of recurrence events within each category. P-values were calculated using Chi-squared and Fisher’s exact tests to compare the number of patients in each group. (**C**) Representative images of NGFR expression in tumor of a non-recurrent (left) and a local recurrent patient (right). Scale-bars: 100 μm.

**Table 1.**
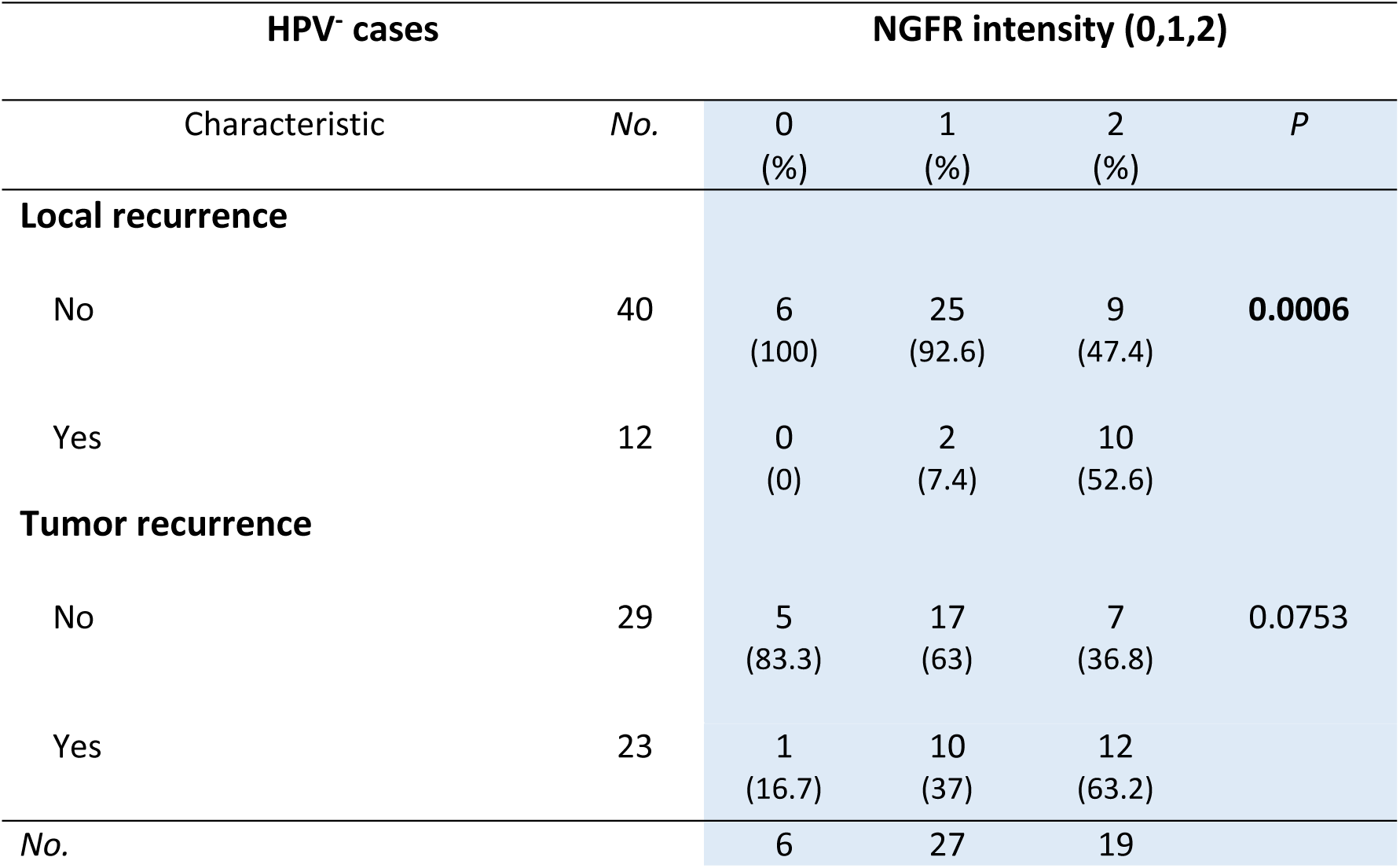
Correlations of NGFR expression by intensity and recurrence in HPV^-^ HNSCC primary tumors.

| HPV<sup>-</sup> cases |  | NGFR intensity (0,1,2) |  |  |  |
| --- | --- | --- | --- | --- | --- |
| Characteristic | No. | 0<br>(%) | 1<br>(%) | 2<br>(%) | *P* |
| Local recurrence |  |  |  |  |  |
| No | 40 | 6<br>(100) | 25<br>(92.6) | 9<br>(47.4) | 0.0006 |
| Yes | 12 | 0<br>(0) | 2<br>(7.4) | 10<br>(52.6) |  |
| Tumor recurrence |  |  |  |  |  |
| No | 29 | 5<br>(83.3) | 17<br>(63) | 7<br>(36.8) | 0.0753 |
| Yes | 23 | 1<br>(16.7) | 10<br>(37) | 12<br>(63.2) |  |
| No. |  | 6 | 27 | 19 |  |

Interestingly, the impact of NGFR on tumor recurrence is specifically associated with its expression intensity, as no significant associations were found when considering staining or tissue distribution patterns (**Supplementary Table 3**). Moreover, the NGFR combined score, which is predictive of survival, did not improve treatment response stratification compared with NGFR intensity alone (**Supplementary Table 3**). We also assessed whether NGFR expression correlated with clinical characteristics. These included general factors such as age and sex; risk factors like smoking and alcohol use; tumor location; TNM staging; histological differentiation; and treatment-related variables such as chemotherapy regimen. Notably, none of these variables showed a significant association with NGFR intensity (**Supplementary Table 4**). Altogether, these findings support a potential role for NGFR in mediating tumor recurrence following RT/CRT, suggesting that NGFR contributes to resistance in HNSCC.

### NGFR is upregulated in Detroit 562 CDDP-resistant model

Given that the prognostic impact of NGFR in our cohort was strongest in HPV-negative oropharyngeal tumors, particularly in patients treated with cisplatin-based chemo-radiotherapy, we next asked whether NGFR upregulation was directly linked to CDDP resistance in experimental models. We therefore focused on CDDP-resistant derivatives of a human oropharyngeal cell line, Detroit 562, which had been generated CDDP-resistant through continuous in vitro exposure to progressively increasing concentrations of the drug [26, 27]. Importantly, *in vivo* analysis of subcutaneous tumors generated by injecting Detroit 562 parental (Detroit 562-P) or CDDP-resistant (Detroit 562-R) cells into immunodeficient nude mice revealed an upregulation of NGFR expression in tumors derived from the resistant line (**Fig. 5A, B**). Consistent with these findings, immunoblot analysis of NGFR in Detroit 562-P and Detroit 562-R cells showed that NGFR is upregulated in the CDDP-resistant model compared with the parental counterpart (**Fig. 5C, D**).

**Figure 5.**
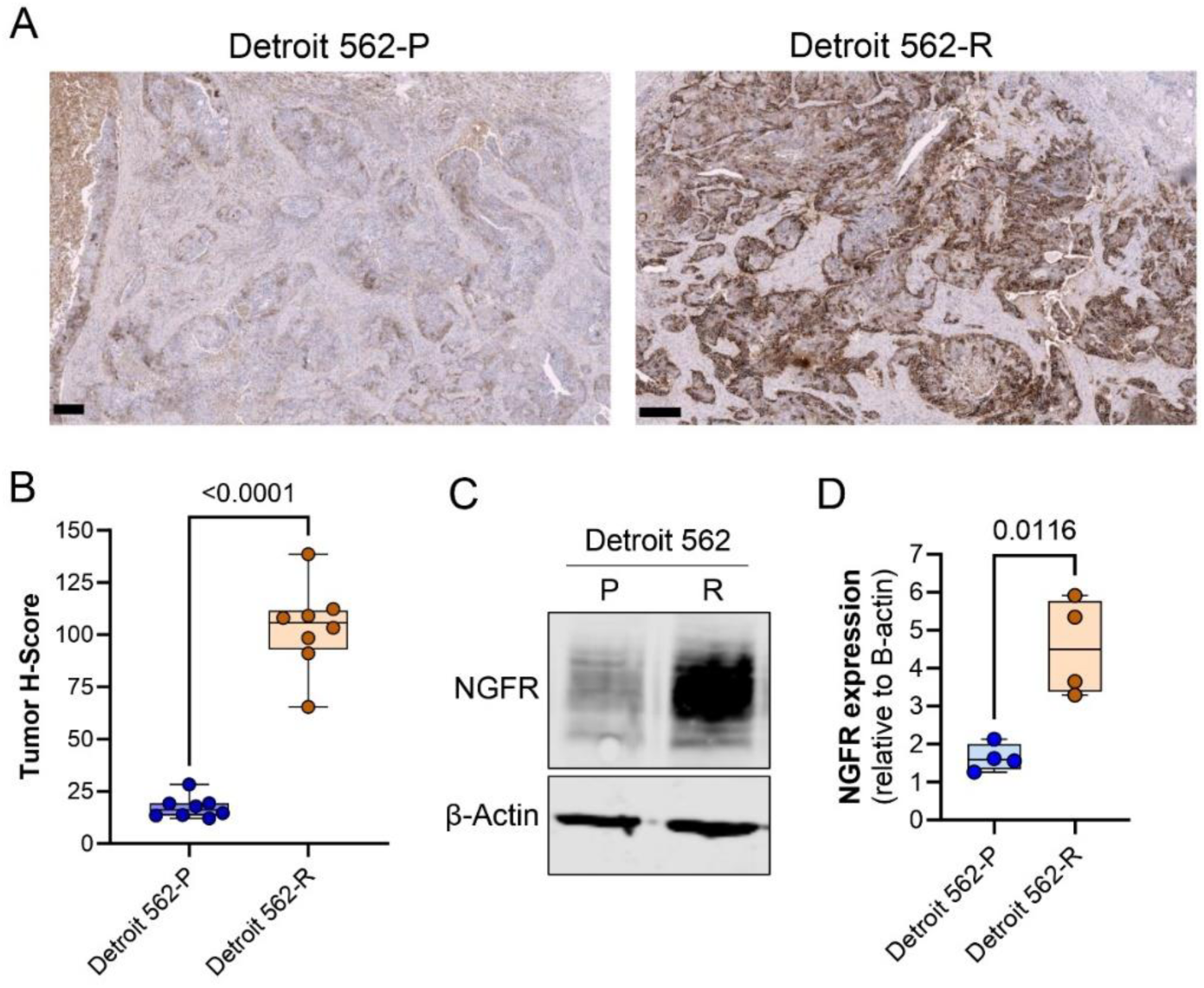
NGFR is upregulated in human HNSCC CDDP-resistant models. (**A, B**) NGFR expression in vivo in Detroit 562-derived tumors measured by immunohistochemistry: representative images (**A**) and quantification (**B**) (n = 8 mice per group). Tumor H-scores were calculated using QuPath (v0.5.1) by first segmenting tumor and stromal regions via machine learning. Staining intensity was categorized as negative (0), low (1), medium (2), or high (3), and the H-score was computed as: (1 × %low) + (2 × %medium) + (3 × %high). P-values for both the immunoblot analysis and tumor staining were calculated using unpaired t-tests. Scale bars: 250 μm. (**C, D**) NGFR expression in vitro in the oropharyngeal Detroit 562 parental (P) and CDDP-resistant (R) cell lines. Representative immunoblot (**C**) and quantification from four independent protein extractions (**D**) are shown.

### NGFR-KO restores CDDP-sensitivity *in vivo*

To evaluate whether NGFR contributes to CDDP resistance, we generated an NGFR-*knockout* (NGFR-KO) model in the Detroit 562-R cell line using CRISPR-Cas9 technology. First, we confirmed the successful depletion of NGFR by immunoblotting (**Fig. 6A**). We then analyzed the effect on primary tumor growth by injecting immunodeficient mice with either the NGFR-KO line or a control line transduced with a non-targeting gRNA (CTL), followed by weekly CDDP treatment (6 mg/kg) (**Fig. 6B**). We found that tumors derived from the Detroit 562-R CTL group were resistant to CDDP, showing no significant impact on mouse survival (log-rank *p* = 0.5933) (**Fig. 6C**) or tumor growth (**Fig. 6D**). In contrast, NGFR KO restored sensitivity to CDDP, resulting in significantly improved survival (log-rank *p* = 0.0071) (**Fig. 6E**) and reduced tumor growth (**Fig. 6F**). These findings suggest that NGFR contributes functionally to CDDP resistance, beyond its role as a biomarker.

**Figure 6.**
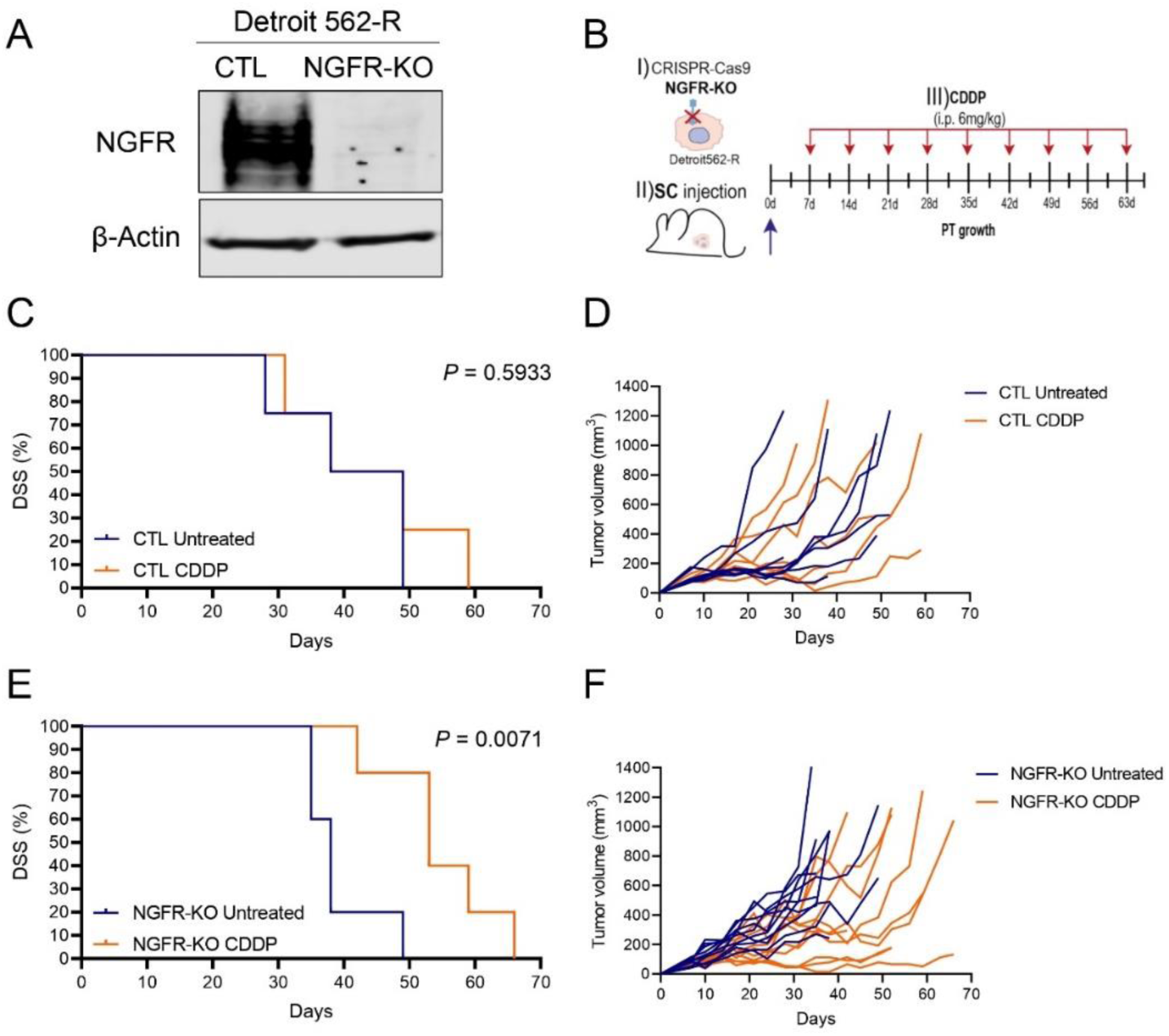
NGFR knockout restores cisplatin sensitivity in vivo in Detroit 562-R–derived tumors. (**A**) Validation of NGFR-KO by immunoblot. (**B**) Schematic representation of the experimental strategy to evaluate the functional role of NGFR in CDDP resistance: (I) we performed an NGFR-KO in the CDDP-resistant Detroit 562-R cell line, characterized by high NGFR expression, using CRISPR–Cas9; (II) NGFR-KO and CTL (empty gRNA) cells were injected subcutaneously into immunodeficient nude mice; (III) once tumors were visible ten days after tumor injection, mice were treated weekly with CDDP (6 mg/kg). (**C, D**) Kaplan–Meier survival curves (**C**) and tumor growth curves (**D**) showing CDDP resistance in the Detroit 562-R CTL model. (E, F) Kaplan–Meier survival curves (**E**) and tumor growth curves (**F**) showing restored CDDP sensitivity in the NGFR-KO model. Sample size: n = 5 mice per group (2 tumors per mouse, 10 tumors/group). P-values for survival analysis were calculated using the log-rank test.

## Discussion

Resistance to standard-of-care RT/CRT remains one of the most critical challenges in the management of HPV-negative HNSCC [5]. Despite advances in radiotherapy techniques and the incorporation of concurrent chemotherapy, a significant number of patients with HPV-negative HNSCC relapse after treatment [5]. In this study, we identify NGFR as a new biomarker of RT/CRT resistance and poor prognosis in HPV-negative HNSCC. Through the analysis of a HNSCC patient cohort, we observed that NGFR is associated with therapeutic resistance. Additionally, we found that NGFR is upregulated in Detroit 562 CDDP-resistant tumor cells and that its genetic depletion restores treatment sensitivity. These findings highlight that NGFR is not only a predictor of recurrence, but also an active driver of therapy resistance, suggesting its potential as both a prognostic marker and a therapeutic target in HNSCC.

Characterization of NGFR expression in the patient cohort showed significant heterogeneity between patients. We observed that NGFR expression ranged from completely negative to strongly positive tumors. Most tumors exhibited low or moderate expression levels, and a subset had very high NGFR expression intensity. Beyond expression intensity, we also described two different staining patterns: in some cases, the expression was limited to the tumor-stroma interface; in other cases, it was diffusely distributed throughout the tumor. Furthermore, we found that in some patients the expression was focal in specific regions of the tumor biopsy, while in others it was homogeneous involving a larger proportion of tumor cells. This homogeneous distribution was often associated with the diffuse staining pattern. Importantly, high NGFR expression correlated with poorer prognosis and reduced patient survival. These findings are consistent with previous reports [23, 25]. In the study by Imai and collaborators, NGFR expression was evaluated in a cohort of 83 hypopharynx tumors, showing variable intensities among patients and demonstrating that NGFR high expression correlated with worse survival [23]. Similarly, another study in oral tumors reported the same correlation between NGFR high expression and poor survival [25]. Interestingly, Søland and collaborators analyzed 53 early-stage (T1-T2N0M0) patients with oral cavity tumors and found that not only a high proportion of NGFR^+^ tumor cells correlated with poor prognosis but also that these tumor cells were distributed in different patterns in the tumor mass or in the invasive fronts [22]. Our observation that NGFR is expressed in some cases in the tumor-stroma interface shows that it could be directly involved in invasive processes as described in this paper. We found that a diffuse pattern of expression that correlates with high numbers of NGFR^+^ tumor cells might have an additive effect with high intensity in determining worse patient survival.

Different studies have characterized the prognostic relevance of NGFR in HNSCC. However, the current knowledge of NGFR in therapy resistance in HNSCC remains limited. In other types of cancer, NGFR expression in tumor cells identifies a multidrug-resistant phenotype. Melanoma is the most studied tumor type, where NGFR^+^ cancer cells are associated with resistance to immunotherapy [28, 29], chemotherapy [20] and targeted therapies [16–19]. Other studies in different tumor types such as breast and esophageal cancer show that NGFR expression is associated with resistance to chemotherapy [30, 31]. One of the few studies that addresses resistance to chemotherapy in HNSCC shows that NGFR⁺ tumor cells upregulate CDDP-resistance–related genes (i.e., ABC transporters) [23]. Another study shows that the neurotrophin axis is relevant for resistance to other treatments such as EGFR inhibitors in HNSCC [32].

Furthermore, our *in vivo* data show that NGFR is not a mere biomarker of resistance but plays a direct and functional role. KO of NGFR in CDDP-resistant cells restored sensitivity to cisplatin in immunodeficient mice, significantly reducing tumor growth and prolonging survival. This result suggests that NGFR inhibition could represent a therapeutic strategy to to overcome RT/CRT resistance in HNSCC. In other tumors, the potential of NGFR inhibitors has demonstrated antitumor effects. For example, small molecules targeting the transmembrane domain of NGFR or a short β-amyloid-derived agonist peptide trigger cell death and slow melanoma tumor growth [33, 34]. Similarly, anti-NGFR antibodies have demonstrated efficacy as monotherapies targeting CSCs [35], or when combined with α-CD47 [36], they exhibit antitumor effects. In addition, strategies that indirectly reduce NGFR expression, such as the use of the heat shock protein inhibitor HSP90 [28], MNK1/2 inhibitors [37], or Ranolazine [38], have resulted in enhanced antitumor immune responses. In addition, we have previously shown that treatment with NGFR inhibitor THX-B [15, 39, 40] reduced melanoma lymph node metastasis [15], although the precise underlying mechanism remains unknown. Despite the growing evidence in other cancers, pharmacologic targeting of NGFR in HNSCC remains largely unexplored. Most functional studies to date have relied on the isolation of NGFR⁺ populations [23] or forced overexpression models [24]. Only one study has evaluated direct NGFR inhibition using a blocking antibody, which effectively abolished the pro-invasive and stem-like phenotype conferred by NGFR [25]. To our knowledge, no studies beyond ours have assessed the impact of NGFR depletion in the context of cisplatin treatment. Further investigation is needed to evaluate the therapeutic potential of NGFR inhibitors, particularly in combination therapy settings.

In conclusion, our findings position NGFR as a new player of therapy resistance in HPV-negative HNSCC. By integrating a patient RT/CRT-treated cohort with *in vivo* functional experiments, we demonstrate that NGFR not only marks a subset of aggressive tumors but also actively contributes to treatment failure. Given its dual role as a prognostic biomarker and therapeutic target, NGFR is a promising candidate for patient stratification and for the development of targeted therapies to overcome RT/CRT resistance. Our work pave the way for future studies to develop NGFR-targeted strategies in HNSCC, as well as elucidating the underlying molecular mechanisms by which NGFR promotes RT/CRT resistance.

## Materials and Methods

### Patient studies

The final cohort included 52 patients with complete clinical, pathological, and follow-up data and with NGFR assessable staining by a pathologist. The majority of cases were oropharyngeal (n = 46), including tumors from several anatomical sub-sites: tonsil (n = 13), base of tongue (n = 24), soft palate (n = 3), and posterior pharyngeal wall (n = 6). A smaller subset of tumors originated in the hypopharynx (n = 6). Tumor staging was diverse, with representation across early and advanced stages: T1 (n = 5), T2 (n = 19), T3 (n = 20), and T4A (n = 8). All patients were treated with radiotherapy (RT). Most also received chemotherapy, primarily cisplatin (n = 32), while others were treated with carboplatin (n = 2) or cetuximab (n = 10). A subset of patients (n = 8) received radiotherapy alone without additional systemic treatment. Comprehensive follow-up data were collected, including recurrence status, survival outcomes, and disease progression. The main clinical characteristics for each patient are summarized in **Supplementary Table 1.**

### Cell lines

For studies investigating the relevance of NGFR in cisplatin (CDDP)-resistant models, we used the Detroit 562-P/R cell lines, derived from the human pharyngeal carcinoma cell line Detroit 562 (RRID:CVCL_1171) developed through *in vitro* exposure of parental Detroit 562 cells to progressively increasing concentrations of cisplatin [26, 27]. We maintained these cell lines in Eagle’s minimum essential medium (EMEM) (Sigma Aldrich) supplemented with 10% fetal bovine serum (Thermo Fisher Scientific), gentamicin (Sigma Aldrich; 0.2 mg/ml) and sodium pyruvate (Sigma Aldrich; 10 mM) in a humidified atmosphere with 5% CO₂ and 85% humidity at 37°C. Detroit-562 cell line used in this study was authenticated by short tandem repeat (STR) profiling and were routinely tested to confirm the absence of Mycoplasma contamination.

### Immunohistochemistry

Tumor sections were stained with an anti-NGFR antibody ([NGFR5], Abcam; ab3125) using the Dako/Agilent Autostainer Link 48. Slides were subsequently scanned using an Axio Scan.Z1 to facilitate expression analysis. A pathologist evaluated NGFR expression intensity and staining patterns in the HNSCC cohort. Among the 52 patients analyzed, NGFR expression was categorized into three intensity levels: negative (n = 6), low (n = 27), and high (n = 19). Two distinct staining patterns were also identified: peripheral, restricted to the outer tumor edge in contact with the surrounding stroma (n = 11), and diffuse, with expression distributed throughout the tumor without a defined pattern (n = 34). Additionally, we identified two tissue distribution patterns: focal, limited to a specific area within the tumor (n = 13), and homogeneous, with consistent expression across the tumor mass (n = 33). The NGFR expression intensity and patterns of each patient together with the clinical characteristics are summarized in the **Supplementary Table 4**.

### Survival and recurrence analysis

We employed Kaplan–Meier analysis for survival analysis, with p-values determined using the log-rank test. We analyzed recurrence-free survival (RFS) to assess associations with treatment resistance, while disease-specific survival (DSS) was used to evaluate outcomes specific to the disease. We conducted the survival analysis based on NGFR expression intensity alone (categorized as 0 = negative; 1 = low; 2 = high) and by combining intensity and expression patterns, represented as (0: negative and low; 1: high) + (1: peripheral; 2: diffuse).

To evaluate whether the prognostic effect of NGFR was an independent prognostic marker, we performed multivariable Cox proportional hazards regression analyses for both RFS and DSS. We fitted separate models in three populations: (i) all HPV-negative HNSCC cases, (ii) HPV-negative oropharyngeal tumors, and (iii) HPV-negative oropharyngeal tumors treated with CDDP. NGFR intensity was modeled as a binary covariate (high vs negative/low, grouping categories 0–1 vs 2), and the combined NGFR score was modeled as a three-level variable with negative/low expression as the reference and high/peripheral and high/diffuse expression entered as separate indicator covariates. All models were adjusted for T stage (T3–4 vs T1–2), nodal status (N+ vs N0), and histological grade (poorly differentiated vs well/moderately differentiated). Hazard ratios and 95% confidence intervals were derived from the Cox model coefficients, and p-values were obtained using Wald tests (**Supplementary Table 2**). We analyzed the association between NGFR expression intensity and patterns with various clinical characteristics (including local and tumor recurrence) using chi-squared and Fisher’s exact tests for comparisons between categorical variables, and unpaired t-tests for comparisons involving continuous variables (**Supplementary Table 3**).

### Immunoblotting

To evaluate NGFR expression levels by immunoblotting, cells were lysed in 1× RIPA buffer containing 0.1% SDS, 0.5% sodium deoxycholate, 1% NP-40 (Igepal-CA-630), 150 mM NaCl, 50 mM Tris-HCl (pH 8), and supplemented with protease and phosphatase inhibitors (Sigma Aldrich). For Western blot analysis the primary antibodies used included NGFR (Abcam, ab52987; 1:1000) and β-actin as a loading control (Santa Cruz, sc-47778; 1:10,000). As secondary antibodies we used fluorescently labeled secondary antibodies IRDye® 800CW anti-rabbit (1:5000, #926-32211, LI-COR) and Alexa Fluor™ 680 anti-mouse (1:5000, #A21057, LI-COR).

Signals were detected using the Odyssey CLx system (LI-COR). The intensities of immunoreactive bands were quantified by densitometry using ImageJ software (ImageJ, RRID:SCR_003070).

### Animal experiments

Athymic nude mice (strain: Athymic Nude-Foxn1nu/nu) were purchased from Envigo. We injected 10⁶ cells from Detroit 562-P/-R cell lines subcutaneously into both flanks of each mouse in a 1:1 Matrigel/PBS mixture. For CDDP treatment, a dose of 6 mg/kg (Casa commercial) was administered once per week via intraperitoneal injection. Tumor growth was monitored twice a week using caliper measurements. Toxicological effects and/or weight loss associated with CDDP treatment were monitored, with early endpoints defined in cases of significant toxicity or weight loss exceeding 10%. The experimental endpoint for survival curves was established when at least one tumor from either flank reached a volume of 1 cm³.

To analyze NGFR expression in IHC-stained Detroit 562-P/-R tumors, we used QuPath (version 0.5.1). First, we configured color deconvolution stains based on hematoxylin and NGFR expression. Then, we performed cell detection to identify individual cells based on hematoxylin staining, followed by positive cell detection to detect NGFR⁺ cells. To determine the varying intensity of NGFR expression, we applied thresholds to classify cells as negative (threshold < 0.2), low or 1+ (threshold > 0.2 and < 0.4), medium or 2+ (threshold > 0.4 and < 0.6), and high or 3+ (threshold > 0.6). Based on these categories, we calculated a single value named H-score using the formula: 1 × percentage of low + 2 × percentage of medium + 3 × percentage of high cells. To ensure the analysis was restricted to tumor cells and excluded stromal regions, we trained an object classifier named “tumor-stroma” by manually annotating representative tumor and stromal areas. In addition, we created a pixel classifier to facilitate automated annotation of the entire tumor area. Finally, we executed a custom script to quantify NGFR expression levels specifically within tumor cells.

## Acknowledgements

We thank the clinical and pathology teams at Hospital Universitario Central de Asturias (HUCA) for their support in patient management and sample collection, and the animal facility staff of the Spanish National Cancer Center (CNIO) for care of the experimental animals. We are also grateful to our colleagues for helpful discussions and technical assistance in this work.

We want to particularly acknowledge for its collaboration the Principado de Asturias BioBank (PT20/00161 and PT23/00077), financed jointly by Servicio de Salud del Principado de Asturias, Instituto de Salud Carlos III and Fundación Bancaria Cajastur and part of the the Spanish National Biobanks Network.

## Funding

This work was supported by the Agencia Estatal de Investigación, Spain (grants PID2020-118558RB-I00 and PDC2021-121102-I00) to H.P. J.G.-A. was supported by Fundación Científica de la Asociación Española Contra el Cáncer (Predoctoral AECC 2021, PRDMA21602GARC). M.E.LL. has received funding from Fondo de Investigación Sanitaria (FIS) project PI24/00630. J.M.G.-P. and J.P.R. received funding from the Instituto de Salud Carlos III (ISCIII) through the project grants PI22/00167, PI25/00107 and CIBERONC (CB16/12/00390) and were co-funded by the European Union. J.M.G.-P. and J.P.R. are funded by the Government of the Principality of Asturias through the Agency for Science, Business Competitiveness and Innovation of the Principality of Asturias and co-financed by the European Union (IDE/2024/000778). M.A.-F. was supported by grant CNS2023-1444-73 funded by MCIU/AEI/10.13039/501100011033 and “EuropeanUnionNextGenerationEU/PRTR” and by the Scientific Foundation of the Spanish Association against Cancer (LABAE235202ALVA).

## Conflict of Interest Statement

The authors declare that they have no conflicts of interest related to this work.

## Ethics Statement

The study involving human tissue samples was conducted in accordance with the Declaration of Helsinki and applicable local regulations. The use of human specimens and associated clinical data was approved by the Ethical and Scientific Committees of the Hospital Universitario Central de Asturias and the Regional Ethics Committee from the Principality of Asturias (CEImPA) (date of approval 9 March 2023; approval number: 2023.018, for the project PI22/00167). Patient informed consent was obtained for all the tissue samples collected by the Biobank of Principado de Asturias (PT20/00161 and PT23/00077).

All animal procedures were carried out in compliance with institutional and national guidelines for the care and use of laboratory animals. The experimental protocols were approved by the corresponding institutional animal ethics committee (Spanish National Cancer Center, CNIO; PROX029.5/22).

## Data Availability Statement

All data supporting the findings of this study are available within the article and its supplementary material. Additional raw data are available from the corresponding author upon reasonable request.

## Abbreviations

CDDP: cisplatin
CRT: chemoradiotherapy
HNSCC: head and neck squamous cell carcinoma
NGFR: nerve growth factor receptor.

## Supplementary Material

**Supplementary Figure 1.**
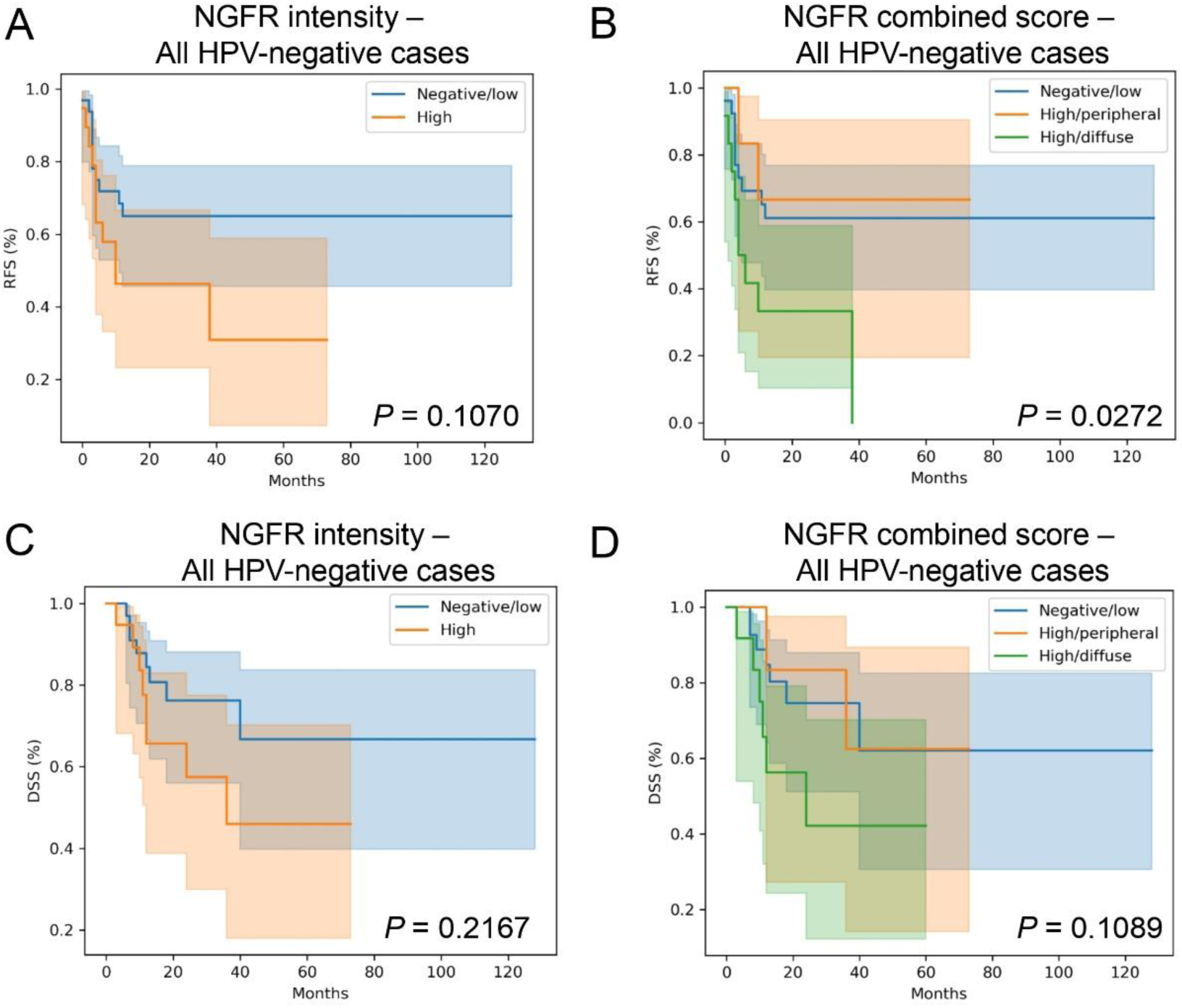
High NGFR expression is associated with poor prognosis in HPV-negative HNSCC. (**A, B**) Kaplan–Meier curves for recurrence-free survival (RFS) in all HPV-negative cases, stratified by NGFR intensity (**A**) or the combined NGFR score (**B**). (**C, D**) Kaplan–Meier curves for disease-specific survival (DSS) in all HPV-negative cases, stratified by NGFR intensity (C) or the combined NGFR score (**D**). NGFR intensity was stratified as high versus negative/low expression. The combined NGFR score was calculated as the product of intensity (0 = negative/low, 1 = high) and staining pattern (1 = peripheral, 2 = diffuse). P-values were calculated using the log-rank test. In panels **B** and **D**, comparisons are shown between negative/low and high/diffuse groups.

---

**Supplementary Table 1.** Clinical characteristics of the HNSCC HPV^-^ human cohort.

| Variable | Category | n | % |
| --- | --- | --- | --- |
| Age (years) | Median [IQR] | 61 [57–66] |  |
| Gender | Male | 48 | 92.3 |
|  | Female | 4 | 7.7 |
| Location | Tonsil | 13 | 25 |
|  | Base of tongue | 24 | 46.2 |
|  | Soft palate | 3 | 5.8 |
|  | Posterior pharyngeal wall | 6 | 11.5 |
|  | Hypopharynx | 6 | 11.5 |
| T classification | T1 | 5 | 9.6 |
|  | T2 | 19 | 36.5 |
|  | T3 | 20 | 38.5 |
|  | T4a | 8 | 15.4 |
| N | 0 | 16 | 30.8 |
|  | 1 | 7 | 13.5 |
|  | 2A | 12 | 23.1 |
|  | 2B | 8 | 15.4 |
|  | 2C | 4 | 7.7 |
|  | 3A | 1 | 1.9 |
|  | 3B | 4 | 7.7 |
| M | 0 | 51 | 98.1 |
|  | 1 | 1 | 1.9 |
| G diff | Well | 11 | 21.2 |
|  | Moderated | 22 | 42.3 |
|  | Poorly | 9 | 17.3 |
|  | Missing | 10 | 19.2 |
| CT | No | 5 | 9.6 |
|  | Yes | 44 | 84.6 |
|  | Missing | 3 | 5.8 |
| CT type | CBDCA | 2 | 3.8 |
|  | CTX | 10 | 19.2 |
|  | CDDP | 32 | 61.5 |
|  | Missing | 8 | 15.4 |

**Supplementary Table 2.** Cox proportional hazards multivariate analysis based on NGFR expression intensity and combined score.

| Endpoint | Subgroup | NGFR model | HR | CI low | CI high | P value | N |
| --- | --- | --- | --- | --- | --- | --- | --- |
| RFS | All HPV-negative | Intensity | 1.943 | 0.84 | 4.494 | 0.12 | 51 |
|  | HPV-negative |  |  |  |  |  |  |
| RFS | oropharynx | Intensity | 2.557 | 1.032 | 6.331 | **0.042** | 46 |
|  | HPV-negative |  |  |  |  |  |  |
| RFS | oropharynx CDDP | Intensity | 3.305 | 1.151 | 9.497 | **0.026** | 30 |
| DSS | All HPV-negative | Intensity | 1.842 | 0.69 | 4.917 | 0.222 | 52 |
|  | HPV-negative |  |  |  |  |  |  |
| DSS | oropharynx | Intensity | 2.347 | 0.755 | 7.297 | 0.141 | 47 |
|  | HPV-negative |  |  |  |  |  |  |
| DSS | oropharynx CDDP | Intensity | 2.553 | 0.636 | 10.245 | 0.186 | 30 |
| RFS | All HPV-negative | Combined | 0.872 | 0.193 | 3.935 | 0.858 | 51 |
| RFS | All HPV-negative | Combined | 3.098 | 1.276 | 7.526 | **0.013** | 51 |
|  | HPV-negative |  |  |  |  |  |  |
| RFS | oropharynx | Combined | 1.266 | 0.273 | 5.874 | 0.763 | 46 |
|  | HPV-negative |  |  |  |  |  |  |
| RFS | oropharynx | Combined | 4.037 | 1.539 | 10.587 | **0.005** | 46 |
|  | HPV-negative |  |  |  |  |  |  |
| RFS | oropharynx CDDP | Combined | 2.135 | 0.44 | 10.352 | 0.346 | 30 |
|  | HPV-negative |  |  |  |  |  |  |
| RFS | oropharynx CDDP | Combined | 4.232 | 1.333 | 13.443 | **0.014** | 30 |
| DSS | All HPV-negative | Combined | 0.989 | 0.208 | 4.708 | 0.989 | 52 |
| DSS | All HPV-negative | Combined | 2.633 | 0.905 | 7.663 | 0.076 | 52 |
|  | HPV-negative |  |  |  |  |  |  |
| DSS | oropharynx | Combined | 0.837 | 0.1 | 7.002 | 0.87 | 47 |
|  | HPV-negative |  |  |  |  |  |  |
| DSS | oropharynx | Combined | 3.737 | 1.127 | 12.391 | **0.031** | 47 |
|  | HPV-negative |  |  |  |  |  |  |
| DSS | oropharynx CDDP | Combined | 1.466 | 0.162 | 13.311 | 0.734 | 30 |
|  | HPV-negative |  |  |  |  |  |  |
| DSS | oropharynx CDDP | Combined | 3.374 | 0.751 | 15.146 | 0.112 | 30 |

**Supplementary Table 3.** Correlations of NGFR intensity, patterns and combined score with local recurrence and tumor recurrence.

| Characteristic | Local recurrence |  | Tumor recurrence |  |
| --- | --- | --- | --- | --- |
|  | No | Yes | No | Yes |
| *Nº of cases* | 40 | 12 | 29 | 23 |
| **NGFR intensity** |  |  |  |  |
| - Negative (0) | 6 | 0 | 5 | 1 |
| - Low (1) | 25 | 2 | 17 | 10 |
| - High (2) | 9 | 10 | 7 | 12 |
| *P* | **0.0006** |  | 0.0753 |  |
| **NGFR staining pattern** |  |  |  |  |
| - Peripheral (1) | 7 | 4 | 5 | 6 |
| - Diffuse (2) | 26 | 8 | 18 | 16 |
| *P* | 0.6567 |  | 0.9324 |  |
| **NGFR tissue pattern** |  |  |  |  |
| - Focal (1) | 11 | 4 | 8 | 7 |
| - Homogeneous (2) | 23 | 8 | 16 | 15 |
| *P* | 1.0000 |  | 1.0000 |  |
| **NGFR combined score** |  |  |  |  |
| - Negative/low (0) | 25 | 2 | 17 | 10 |
| - High/peripheral (1) | 3 | 3 | 3 | 3 |
| - High/diffuse (2) | 5 | 7 | 3 | 9 |
| *P* | **0.0015** |  | 0.0909 |  |

**Supplementary Table 4.**
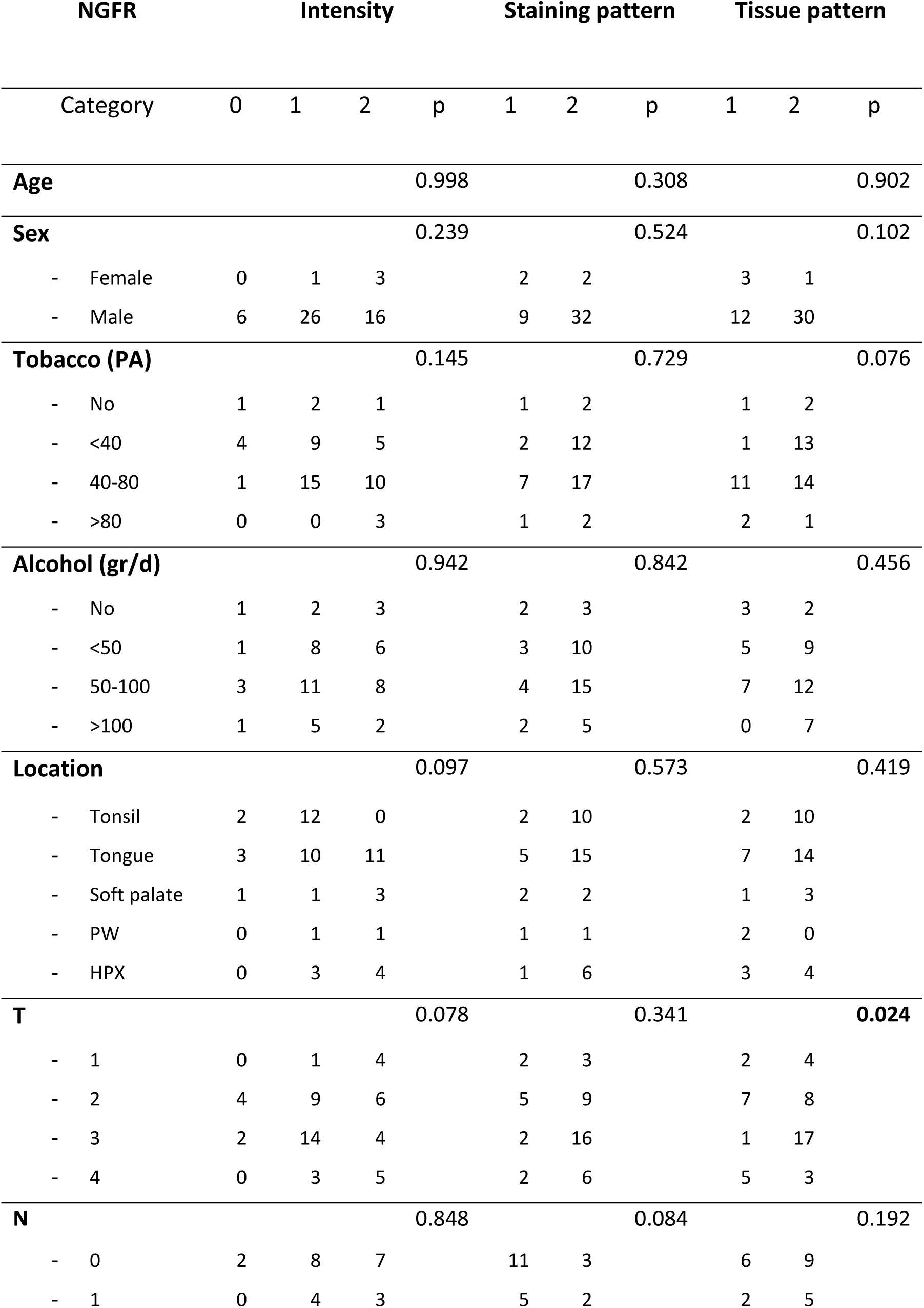
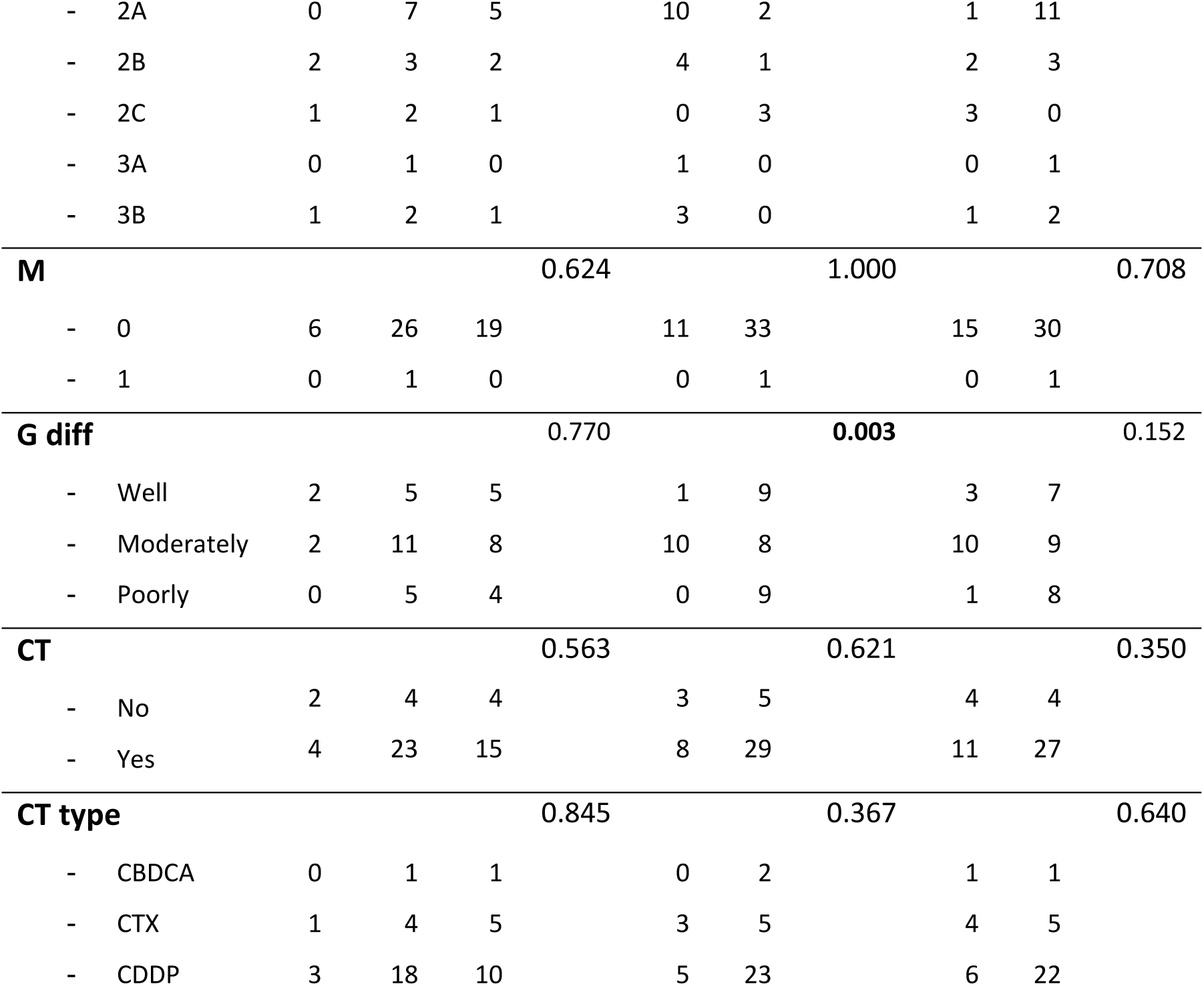
Associations of NGFR expression intensity and patterns with clinical characteristics of the cohort.

## Notes

### Competing Interest Statement

The authors have declared no competing interest.

### Funding Statement

This work was supported by the Agencia Estatal de Investigacion, Spain (grants PID2020-118558RB-I00 and PDC2021-121102-I00) to H.P. J.G.-A. was supported by Foundation of the Spanish Association against Cancer (Predoctoral AECC 2021, PRDMA21602GARC). M.E.LL. has received funding from Fondo de Investigacion Sanitaria (FIS) project PI24/00630. J.M.G.-P. and J.P.R. received funding from the Instituto de Salud Carlos III (ISCIII) through the project grants PI22/00167, PI25/00107 and CIBERONC (CB16/12/00390) and were co-funded by the European Union. J.M.G.-P. and J.P.R. are funded by the Government of the Principado of Asturias through the Agency for Science, Business Competitiveness and Innovation of the Principado of Asturias and co-financed by the European Union (IDE/2024/000778). M.A.-F. was supported by grant CNS2023-1444-73 funded by MCIU/AEI/10.13039/501100011033 and EuropeanUnionNextGenerationEU/PRTR and by the Scientific Foundation of the Spanish Association against Cancer (LABAE235202ALVA).

